# Distal gene regulation mediated by non-coding RNAs contributes to germline risk for breast and prostate cancer

**DOI:** 10.1101/2022.02.08.22270601

**Authors:** Nolan Cole, Paige Lee, Tommer Schwarz, Pan Zhang, Matthew L. Freedman, Alexander Gusev, Sara Lindström, Michael J. Gandal, Bogdan Pasaniuc, Arjun Bhattacharya

## Abstract

Genome-wide association studies (GWAS) have identified numerous genetic loci associated with breast and prostate cancer risk, suggesting that germline genetic dysregulation influences tumorigenesis. However, the biological function underlying many genetic associations is not well-understood. Previous efforts to annotate loci focused on protein-coding genes (pcGenes) largely ignore non-coding RNAs (ncRNAs) which account for most transcriptional output in human cells and can regulate transcription of both pcGenes and other ncRNAs. Though the biological roles of most ncRNAs are not well-defined, many ncRNAs are involved in cancer development. Here, we explore one regulatory hypothesis: ncRNAs as *trans*-acting mediators of gene expression regulation in non-cancerous and tumor breast and prostate tissue. Using germline genetics as a causal anchor, we categorize distal (>1 Megabase) expression quantitative trait loci (eQTLs) of pcGenes significantly mediated by local-eQTLs of ncRNAs (within 1 Megabase). We find over 300 mediating ncRNAs and show the linked pcGenes are enriched for immunoregulatory and cellular organization pathways. By integrating eQTL and cancer GWAS results through colocalization and genetically-regulated expression analyses, we detect overlapping signals in nine known breast cancer loci and one known prostate cancer locus, and multiple novel genetic associations. Our results suggest a strong transcriptional impact of ncRNAs in breast and prostate tissue with implications for cancer etiology. More broadly, our framework can be systematically applied to functional genomic features to characterize genetic variants distally regulating transcription through *trans*-mechanisms.

**SIGNIFICANCE:** This study identifies non-coding RNAs that potentially regulate gene expression in *trans*-pathways and overlap with genetic signals for breast and prostate cancer susceptibility, with implications for interpretation of cancer genome-wide association studies.

## INTRODUCTION

Genome-wide association studies (GWAS) of cancer risk have revealed risk-associated alleles at hundreds of genetic loci, with breast and prostate cancer GWAS yielding the largest number of associations (1, 2). Through integration with transcriptomic and functional genomics datasets, the proposed target genes for many of these risk loci have been found in protein-coding regions of the genome (1–4). However, many risk variants fall in non-coding regions of the genome and, for these variants, identifying the likely biological mechanism is challenging. One proposed mechanism for GWAS-identified risk variants is *trans*-acting pathways: a GWAS variant affects a regulatory feature, like a transcription factor, in proximity, which then affects genes located far away from the GWAS variant. Particularly, one study identified GWAS risk variants for breast cancer that confer *trans*-effects through transcription factors, like *ESR1*, *MYC*, and *KLF4* (5). Another potential mechanism by which GWAS variants in non-coding regions affect risk is mediation of *trans*-acting effects of genetic variants through non-coding RNAs (ncRNAs).

Although ncRNAs do not code for proteins, they account for nearly 60% of transcriptional output in human cells and interact with a complex network of genes, transcripts, and proteins with widespread effects on cell biology (6, 7). Some ncRNAs, like microRNAs (miRNAs), target and degrade mRNA transcripts of specific genes and link to regulatory networks that include multiple ncRNAs and protein-coding genes (pcGenes). These complex interactions between ncRNAs and pcGenes support the hypothesis that ncRNAs have key roles in cellular pathways (6,8,9). Specific ncRNAs have been shown to leave their transcription site and regulate gene expression at genomic regions far from their transcription start site (8, 9). However, regulatory impacts of ncRNAs on transcription of pcGenes are generally uncategorized, especially in a systematic fashion (8). ncRNAs have shown associations with the onset and progression of different cancers, are enriched in multiple tumor types, and are even therapeutic targets, as they act as regulators of genes in important tumorigenic or progressive networks (10). For example, the long ncRNA *XIST* exerts oncogenic and metastatic effects in multiple cancer types (11). Profiling and deep sequencing of ncRNAs have shown that perturbing ncRNA biogenesis affects amplification, deletion, and normal epigenetic and transcriptional regulation (10,12–14); accordingly, ncRNAs can act as oncogenes or antagonize tumor suppressors.

However, as most ncRNA mechanisms in cancer tumorigenesis or progression have been categorized on a case-by-case basis (13, 14), mechanistic impacts of ncRNAs have not been explored systematically. Bioinformatics analyses that leverage high-throughput genomics have investigated the role of ncRNAs through computational target prediction or differential expression analyses. Although these computational methods have elucidated potential roles of ncRNAs in cancer, they have limitations, including computational feasibility and functional translation of sequence similarity methods (15) and reverse causality for differential expression analyses (i.e., differential expression more likely reflects consequences of disease) (16).

One systematic approach to identifying potential *trans*-mechanisms of regulation is to use genetic variants as causal anchors. A prevailing thought is that distal expression quantitative trait loci (eQTLs) of genes, where the genetic variant is far away from the gene (more than 1 Megabase, or Mb), are often themselves local-, or *cis*-acting, QTLs of a regulatory feature (17–22). We emphasize that the modifiers “local” and “distal” refer merely to distances in the genome (i.e., within or outside 1 Mb, respectively), whereas *cis*- and *trans*-acting refer to the biological mechanism (i.e., direct or indirect interaction, respectively). Molecular features, like ncRNAs, that have potential *trans*-acting regulatory effects can be identified through mediation analyses, either at variant- or gene-level (18,21,23,24). Not only can these analyses point to distal relationships between ncRNAs and pcGenes, but they can point to genetic variants associated with disease etiology with potential distal effects in the transcriptome.

Here, we systematically map distal-eQTLs of pcGenes that are potentially mediated by local-eQTLs of ncRNAs in non-cancerous and tumor prostate and breast tissue, using data from the Genotype Tissue-Expression (GTEx) Project (25) and The Cancer Genome Atlas (TCGA) (26). We then employ colocalization (27) and genetically-regulated expression analysis (28) to identify overlaps in eQTLs and GWAS signals for both overall and molecular subtype-specific breast (2) and overall prostate (1) cancer risk. In total, our work shows the widespread transcriptomic impact of genetically-mediated portion of ncRNAs and that this impact has key associations with cancer susceptibility. This approach provides a rigorous framework to not only categorize functional hypotheses of distal regulatory effects of ncRNAs but also other regulatory molecular features.

## MATERIALS AND METHODS

A graphical representation of our methods is provided in Supplemental Figure S1.

### Data acquisition and processing

We used pre-processed genotype, transcriptomic, and covariate data for non-cancerous mammary and prostate tissue from the Genotype-Tissue Expression Project (GTEx) v8 (25) and breast and prostate tumor tissue from The Cancer Genome Atlas (TCGA) (26). We included only individuals of European ancestry due to the small sample sizes available for other demographics (*N* = 337 for GTEx breast, *N* = 186 for GTEx prostate, *N* = 437 for TCGA breast, *N* = 349 for TCGA prostate). For both GTEx and TCGA, we only consider SNPs and genes on autosomes, restricted to SNPs with minor allele frequency greater than or equal to 1%, and excluded SNPs that deviated from Hardy-Weinberg equilibrium at *P* < 10^−6^.

We used the BioConductor package *biomaRt* for ENSEMBL gene biotype annotations (29). Using these annotations, we defined pcGenes as those labeled “protein-coding” and non-coding RNAs (ncRNAs) as those labeled otherwise; we exclude transcripts labelled as “pseudogenes”. These annotations included 16,582 pcGenes and 5,650 ncRNAs in GTEx breast, 16,827 pcGenes and 5,862 ncRNAs in GTEx prostate, 21,648 pcGenes and 1,261 ncRNAs in TCGA breast, and 15,773 pcGenes and 548 ncRNAs in TCGA prostate. We considered all provided GTEx covariates: 5 genotype-based principal components (PCs), up to 60 probabilistically-estimated expression residuals (PEER) factors, age, sex, and sequencing platform and protocol (25). For TCGA, we calculated genotype PCs using PLINK v1.93 (30), calculated up to 50 hidden components of expression (HCP) using *Rhcpp* (31, 32), and included the following covariates: age, estrogen receptor subtype, menopausal status, and disease pathological stage. For prostate tumors, we include the following covariates: age, sequencing platform, and protocol.

We integrated eQTL results with GWAS summary statistics for overall and subtype-specific breast and overall prostate cancer risk. We obtained European-ancestry specific overall and subtype-specific GWAS summary statistics for breast cancer risk from the BCAC Consortium (2). We studied 5 intrinsic breast cancer molecular subtypes, defined by combinations of estrogen (ER)-, progesterone (PR)-, and human epidermal growth factor receptor (HER2) and tumor grade (2): Luminal A-like (ER + and/or PR +, HER2-, grade 1 or 2); (2) luminal B-like/HER2-negative (ER + and/or PR +, HER2-, grade 3); (3) luminal B-like/HER2-positive (ER + and/or PR +, HER2 +); (4) HER2-positive/non-luminal (ER-and PR-, HER2+), and (5) TNBC (ER-, PR-, HER2-). We obtained European-ancestry specific GWAS summary statistics for prostate cancer risk from the PRACTICAL Consortium (1).

### eQTL mapping

We used a multiple linear regression model in *MatrixEQTL* to detect local- and distal-eQTLs (33). Here, we define a local-eQTL as a variant within 1 Mb of the gene body and a distal-eQTL as a variant outside the 1 Mb window. eQTLs outside this 1 Mb window are unlikely to have direct effects on the promoters or enhancers of the gene and are more likely to have *trans*-acting mechanisms (17, 18).

To determine a set of covariates that maximizes the number of detected distal-eQTLs for pcGenes, we iterated on eQTL mapping using SNPs on Chromosome 22. For breast tumor tissue, we found that the optimized covariate set for local-eQTL mapping included all of the clinical covariates (age, estrogen receptor subtype, menopause status, and disease pathological stage), the first 3 PCs, and the first 8 HCPs. For prostate tumor tissue, we included age, sequencing platform, protocol as covariates and used 5 genotype PCs and 10 HCPs as the optimized set of covariates (31). For data from GTEx, we used the full set of provided covariates for non-cancerous breast and prostate tissue local- and distal-eQTL mapping. We then run genome-wide eQTL mapping with these optimized sets of covariates.

### Mediation analysis for distal-eQTL mapping

We first identify a testing triplet, consisting of (1) a SNP *s*, (2) a distal pcGene *G* associated with SNP *s* with nominal *P* < 10^−6^, and (3) a set of local ncRNAs *m*_1_, …, *m*_*M*_, all associated with SNP *s* with nominal *P* < 10^−6^. As in previous distal-eQTL studies, we use a liberal P-value threshold to increase the number of testing triplets subjected to rigorous permutation-based mediation analysis (18,21,23). Next, we fit the following two sets of linear regressions for mediation analysis (21, 23):

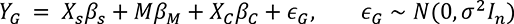

where *Y*_*G*_ is a vector of expression for gene *G*, *X*_*s*_ is a vector of dosages for SNP *s*, *β*_*s*_ is the effect size of SNP *s* on gene *G*, *M* is the expression matrix of *m* ncRNAs, *β*_*M*_ is the effects of the *M* ncRNAs on *Y*_*G*_, *X*_*C*_ is a matrix of covariates, and *ε*_*G*_ is a random error term. The *j*th ncRNA is modeled as

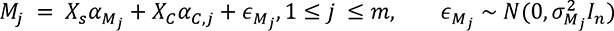

where *M*_*j*_ is the vector of expression for the *j*th ncRNA, α_*Mj*_ is a vector of effects of SNP *s* on mediator *M*_*j*_, *X*_*C*_ is a matrix of covariates, α_*C*,*j*_is a vector of covariate effects on the mediator, and *ε*_*Mj*_ represents a random error term.

We define the total mediation effect (TME) as *TME* = *α_*M*_* ⋅ *β*_*M*_ and the mediation proportion (MP) as *M*P** = min 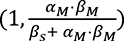. We test *H*_0_ : *TME* = 0 vs. *H*_1_ : *TME* ≠ 0 via permutation testing with 10,000 draws.

### Gene-based association testing (GBAT)

We applied GBAT (24) with modifications to identfy ncRNAs with genetically-regulated expression (GReX) associated with distal pcGenes. First, we removed multi-mapped reads using previously provided annotations (34). Then, we estimated the heritability of ncRNA expression using GCTA v1.93 (35), excluding genes with limited evidence of heritability (P > 0.05). Next, using leave-one-out cross-validation, we constructed the ncRNA GReX using SNPs within 1 Mb using elastic net, LASSO (36) and SuSiE (37), excluding ncRNAs with cross-validation R^2^ < 0.01. We then employed MatrixEQTL to estimate the association between the ncRNA GReX and distal pcGene expression, adjusting for the optimized set of covariates from eQTL mapping (33). Lastly, to identify a set of SNPs that best explains distal ncRNA-pcGene association, we used SuSiE fine-mapping with default parameters to define a 90% credible set (37).

### Colocalization with cancer risk

To identify any potentially overlapping signals between local-eQTLs of ncRNAs, ncRNA-mediated distal-eQTLs of pcGenes, and cancer risk, we employed the Bayesian colocalization method, *coloc* (27). *coloc* estimates the posterior probability that the same SNP explains both the eQTL and the GWAS signal at a given locus. We used standard parameters with default priors (p_1_ = 10^-4^, p_2_ = 10^-4^, and p_12_ = 10^-6^) to estimate the colocalization posterior probability. We considered an eQTL signal to colocalize with a GWAS signal if the posterior probability of colocalization through one SNP (PP.H4 in Giambartolomei et al) was greater than 0.75 (27).

### Genetically-regulated expression analysis of ncRNAs

We identified any cancer associations for the genetically-regulated expression (GReX) of any ncRNAs that showed significant mediation of multiple distal-eQTLs of distant pcGenes. First, using elastic net regression, linear mixed modeling, and SuSiE, we built predictive models of ncRNAs in both GTEx and TCGA data across breast and prostate tissue and select only models with 5-fold cross-validation McNemar’s adjusted R^2^ > 0.01 (28,36–38). We then employed the weighted burden test and permutation test from the FUSION TWAS framework to detect a trait association with the GReX of an ncRNA (28). We define transcriptome-wide significance as P < 2.5 x 10^-6^ and permutation test P < 0.05.

### Data Availability

GTEx v8 data were obtained through dbGAP Study Accession phs000424.v8.p2. TCGA genotype were obtained through dbGAP Study Accession phs000178.v11.p8 and expression and covariate data was obtained from the Broad GDAC Firehose repository (https://gdac.broadinstitute.org). Prostate cancer GWAS summary statistics were obtained from the Prostate Cancer Association Group to Investigate Cancer Associated Alterations in the Genome (PRACTICAL) Consortium: http://practical.icr.ac.uk/blog/wp-content/uploads/uploadedfiles/oncoarray/MetaSummaryData/meta_v3_onco_euro_overall_ChrAll_1_release.zip. Breast cancer GWAS summary statistics were obtained from the Breast Cancer Association Consortium (BCAC): https://bcac.ccge.medschl.cam.ac.uk/bcacdata/oncoarray/oncoarray-and-combined-summary-result/gwas-summary-associations-breast-cancer-risk-2020/. Sample code for this analysis is available at https://github.com/ColetheStatistician/ncRNAInBreastCancer/.

## RESULTS

In this work, we uncover hidden mechanisms contributing to genetic risk for breast and prostate cancer mediated by ncRNAs, systematically exploring one regulatory hypothesis: distal mediation of pcGene expression regulation in non-cancerous and tumor breast and prostate tissue (Figure 1). Specifically, we identify distal-eQTLs of protein-coding genes (pcGenes) that are significantly mediated by ncRNAs local to these distal-eQTLs and assess if they overlap with genetic signal for cancer risk.

**Figure 1:**
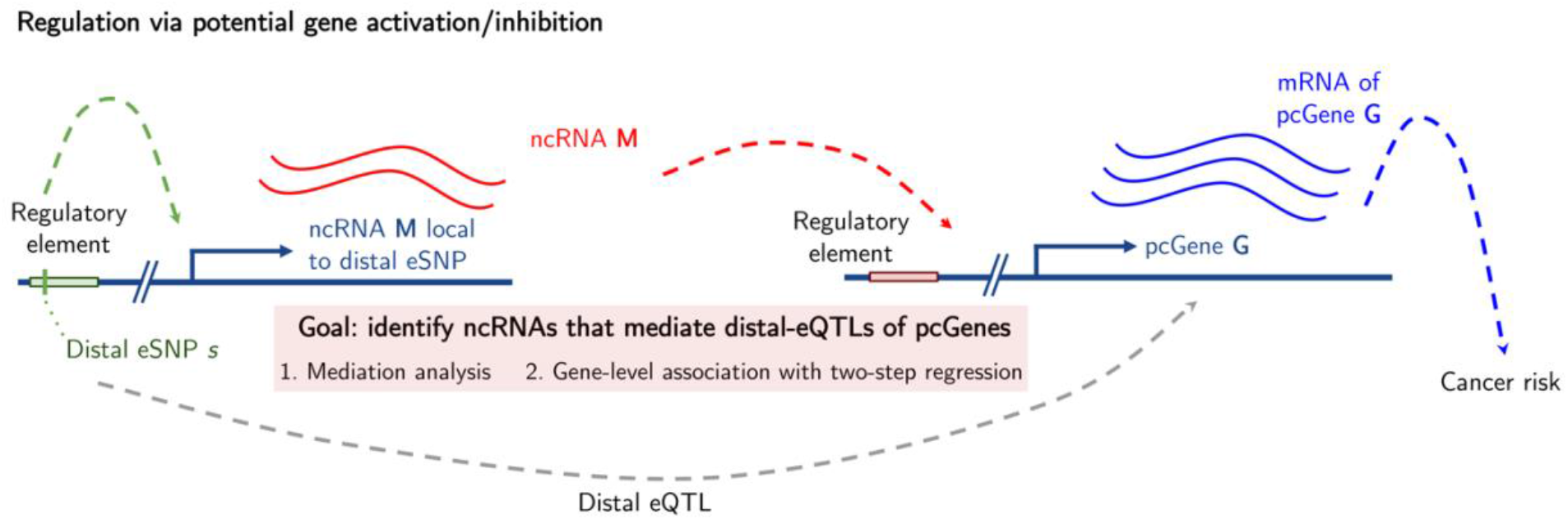
Schematic of mediation analysis to identify ncRNA-mediated distal-eQTLs of protein-coding genes. A SNP s is distal to protein-coding gene G and local to ncRNA M, where M has some distal regulatory effect on G. We use expression quantitative trait locus mapping to identify the local-eQTL between s and M (green dotted line) and the distal-eQTL between s and G (grey dotted line). Using either mediation analysis or gene-level association testing, we estimate the indirect mediation effect of s on G through effects from M (red line). Lastly, we use colocalization and genetically-regulated expression analysis to find any intersecting genetic signal between distal-eQTLs of G and genetic associations with breast and prostate cancer risk.

### Multiple ncRNAs mediate distal-eQTLs in breast and prostate tissue

#### Distal-eQTL mapping through mediation analysis

We conducted distal-eQTL mapping through ncRNA mediation in GTEx (25) and TCGA (26). The number of distal-eQTLs of pcGenes that are significantly mediated by ncRNAs are reported in Table 1 and **Supplemental Table S1-2**. Distributions of TME and MP in tumor tissue showed a larger range than in non-cancerous tissue for both breast and prostate, and median TME and MP were higher in tumor tissue (Supplemental Figure S2).

**Table 1:** Summary of local- and distal-eQTL mapping results across breast and prostate non-cancerous and tumor tissue through mediation analysis.

|  |  | *Breast* |  | *Prostate* |  |
| --- | --- | --- | --- | --- | --- |
|  |  | Non-cancerous | Tumor | Non-cancerous | Tumor |
| **Total eQTLs** |  |  |  |  |  |
|  | *Local* | 22,832 | 1,298 | 12,511 | 6,368 |
|  | *Distal* | 29,512 | 316,097 | 31,782 | 363,587 |
| **Total eGenes** |  |  |  |  |  |
|  | *local-ncRNAs* | 1,113 | 87 | 773 | 60 |
|  | *distal-pcGenes* | 8,849 | 19,376 | 8,711 | 15,560 |
| **SNPs associated in both local and distal eQTLs** |  |  |  |  |  |
|  | *Local* | 1,569 | 601 | 281 | 6,368 |
|  | *Distal* | 1,580 | 5,089 | 264 | 9,259 |
| **Mediated distal-eQTLs** |  |  |  |  |  |
|  | *eSNPs* | 703 | 3,017 | 103 | 425 |
|  | *e-ncRNAs* | 157 | 45 | 22 | 18 |
|  | *e-pcGenes* | 173 | 562 | 24 | 107 |

In non-cancerous breast tissue, pcGenes with cross-chromosomal distal-eQTLs with large ncRNA-mediated effects included *PUM1* and *PCBP1* (Figure 2A), both of which influence tumorigenesis (39–41). In breast tumors, we found *MT4* and *GPRC6A* have large mediated distal effects. *MT4* belongs to the metallothionein family involved in breast cancer carcinogenesis (42, 43), and *GPRC6A* is a part of the androgen receptor signaling pathway (44, 45). In non-cancerous prostate tissue, we detected large distal mediation effects for genes such as *RAF1*, a proto-oncogene (46) and *LGR5*, a gene associated with prostatic regeneration and overexpressed in prostate tumors (47). We also detected a number of olfactory receptors (*OR2T1*, *OR10G8*, *OR10S1*) with large mediated effects in prostate tumor. Olfactory receptors are associated with prostate cancer progression but generally have low expression in prostate tumors (48, 49).

**Figure 2:**
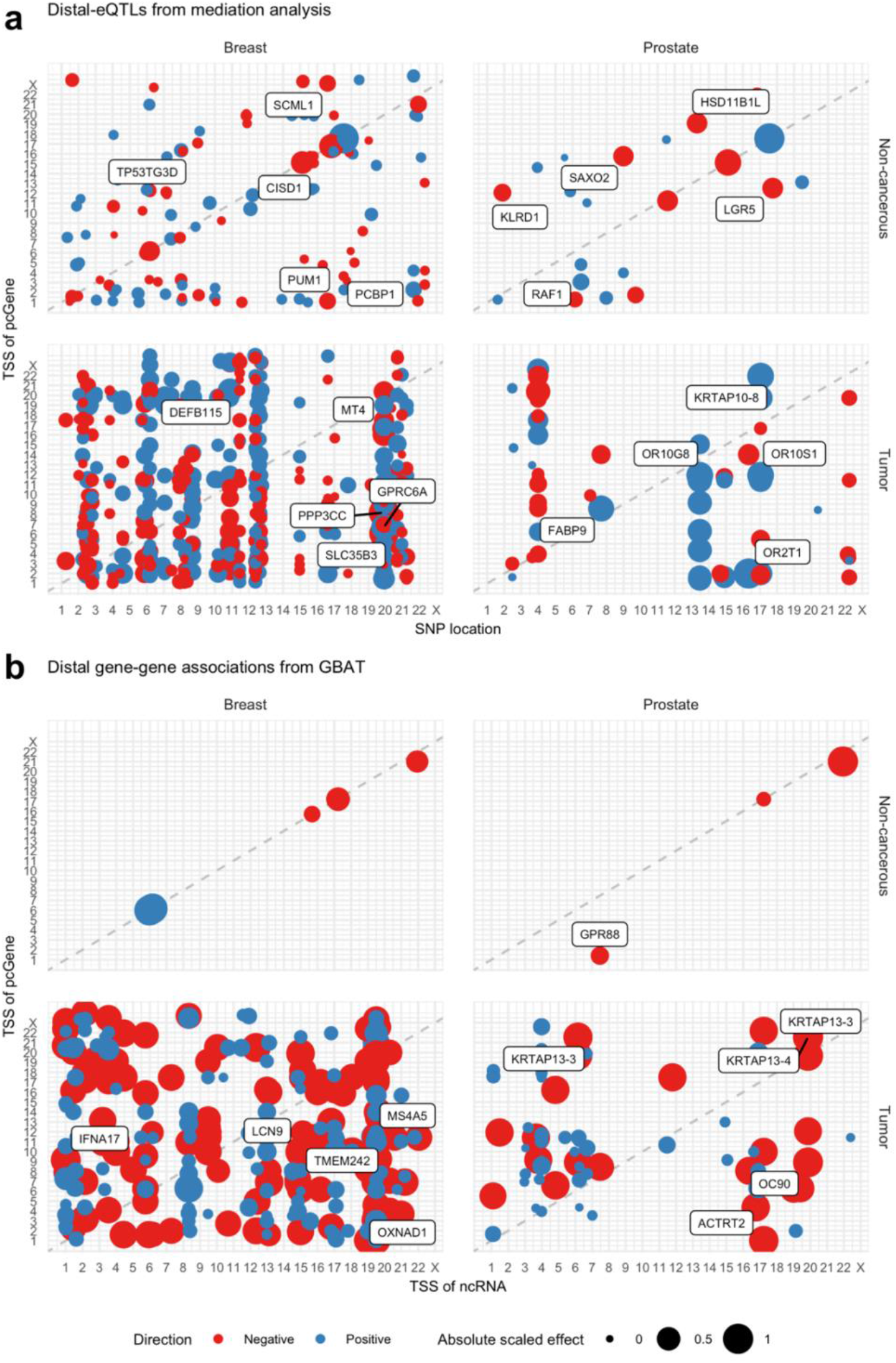
Location plot of distal-eQTL or gene-gene associations across healthy and tumor samples of breast and prostate tissue. **(a)** eSNP (X-axis) position vs. transcription start site (TSS) of pcGene (Y-axis) at FDR-adjusted P < 0.01, sized by absolute scaled TME and colored by direction of effect. **(b)** ncRNA TSS (X-axis) vs. pcGene TSS (Y-axis) at FDR-adjusted P < 0.01, sized by absolute scaled gene-gene effect and colored by direction of effect. Top cross-chromosomal distal-eGenes with largest effects are labeled.

Owing mainly to a larger set of nominally-significant distal-eQTLs in tumor tissues and larger sample sizes in TCGA than GTEx, we found nearly four times as many significantly mediated distal-eQTLs in tumor tissue compared to non-cancerous tissue. Among distal-eQTLs detected in tumor samples, we observed that multiple ncRNAs mediated distal-eQTLs with many different pcGenes (vertical bands in Figure 2A). One such ncRNA in breast tumors is *LINC00301*, showing significant mediation of 1,103 distal-eQTLs across 53 unique SNPs and 53 unique pcGenes. Many of these pcGenes belong to the GAGE protein family, which promotes breast cancer cell invasion and has shown evidence of distal genetic regulation (50, 51). *LINC00301* itself has been implicated in facilitating tumor progression and immune suppression, albeit in lung cancer (52). Another example of such an ncRNA in breast tumors is *miR-548f-4*, a commonly mutated microRNA in multiple cancers (53), mediating more than 300 distal-eQTLs for 16 unique pcGenes, including *GUCA2B*, upregulated in breast cancer metastases (54), and *CYP2C9*, a target of tamoxifen (55). In contrast, only two ncRNAs in non-cancerous breast tissue showed significant mediation of more than 50 distal pcGenes. One of these genes, *FAM106A*, mediated distal-eQTLs of 13 pcGenes, including *BTN3A2*, a prognostic marker for breast cancer (56). In both non-cancerous and tumor breast tissue, three pcGenes (*PRCC1*, *CYP2C9*, and *ATG14*) showed significant mediation through ncRNAs, though the sets of pcGene targets across non-cancerous and tumor tissue are distinct.

In prostate tumors, *LINC02903* showed significant mediation with the most distal-eQTLs (177 eQTLs across 8 pcGenes). These pcGenes include *FABP9*, an upregulated gene in prostate carcinomas with prognostic value (57), and *MTNR1B*, a gene harboring nominal risk variants for prostate cancer (58). Another ncRNA with significant TME for nearly 100 distal-eQTLs across 20 pcGenes was *SDHAP2*. Many of these pcGenes have been implicated in prostate cancer and metastasis pathways, including *TMEM207*, *FADS6*, *MTNR1B, SLC26A8,* and *FGF23* (59–61). Similar to breast tissue, only two ncRNAs showed significant mediation of more than 20 distal eQTLs in prostate tissue: *FBXO30-DT* and *SNHG2*, which has been implicated in tumorigenesis and proliferation in multiple cancers (62–64). A majority (22/26) of distal-eQTLs mediated by *FBXO30-DT* are for *OVCH2*, which has been implicated in prostate risk through GWAS (65, 66). The majority of distal-eQTLs mediated by *SNHG2* are for *RAF1*, a therapeutic target for multiple cancers (46). We did not detect any shared ncRNAs or pcGenes across non-cancerous and tumor prostate tissue.

#### Gene-based distal-eQTL mapping

Next, we conducted gene-level distal eQTL mapping using GBAT (24) to identify ncRNAs that are regulated by multiple weak local genetic effects and may have distal effects on pcGenes; these ncRNAs are likely to be missed by the mediation framework. Comparing to mediated pcGenes from mediation analysis (Table 1), we found a similar order of magnitude of pcGenes with distal associations with ncRNAs using GBAT (Table 2). We again detected far more distal ncRNA-pcGene directional associations in tumor compared to non-cancerous tissue (Table 2**, Supplemental Table S1-2**). In addition, the distribution of ncRNA-pcGene effect sizes is shifted downwards in tumor tissue, compared to non-cancerous tissue, though the number of effect sizes in these distributions are far less for non-cancerous tissue distal associations (Supplemental Figure S2). In breast tumors, we found large distal genetic associations with pcGenes like *OXNAD1*, an RNA-binding protein that is associated with pan-cancer survival rates and involved with tumor invasion and metastasis (66, 67), and *LCN9*, part of the lipocalin family that promotes breast cancer metastasis (68). In prostate tumors, we detected multiple large distal genetic associations with genes in the pro-proliferative keratin-associated protein family (*KRTAP13-3*, *KRTAP13-4*, *KRTAP10-8*) (69) (Figure 2).

**Table 2:** Summary of distal-eQTL mapping results across breast and prostate normal and tumor tissue using GBAT.

|  | *Breast* |  | *Prostate* |  |
| --- | --- | --- | --- | --- |
|  | Normal | Tumor | Normal | Tumor |
| *Total gene-gene associations* | 13 | 1,375 | 7 | 297 |
| *Unique ncRNAs* | 9 | 209 | 4 | 84 |
| *Unique pcGenes* | 10 | 1,127 | 5 | 268 |

Though distal genetic effects of ncRNAs in non-cancerous breast or prostate tissue were sparse, we found three ncRNAs with genetic associations with pcGenes across both breast and prostate tissue: *LINC01678*, *FAM106A*, and *AP001056.1*. These ncRNAs also target the same pcGenes (*LOC102724159*, *CCDC144A*, *GATD3B*, and a paralog to *TRAPPC10*), all with no catalogued functions in cancer.

Again, in tumor-specific gene-gene associations, many ncRNAs had associations with multiple pcGenes, leading to vertical bands in the location plots in Figure 2B. For example, in breast tumors, *LINC000906*, a predicted miRNA sponge in breast tumors (70), was associated with 115 pcGenes. The largest association was with *MS4A5*, whose hypomethylation is shown to be prognostic for multiple cancer types (71, 72). Another ncRNA associated with multiple different pcGenes *is LINC00115*, a known promoter of breast cancer metastasis and progression (73–75). Many of these targets are related to interferons (*IFNA17* and *IFNW1*), immune system cytotoxicity (*RAC2* and *DDB2*), or secretory proteins (*PRH1* and *PRH2*).

In prostate tumors, *FAM138F* showed distal genetic associations with 57 distinct pcGenes, many of which are involved in amino acid activation in prostate cancer (*HDAC2*, *NT5DC1*, *NUS1*, and *PREP*) and protein stability (*PDCD2*, *TCP1*). Additionally *TDRG1*, shown to be associated with progression and metastases in multiple cancers (76, 77), showed multiple distal genetic associations with pcGenes, including the cancer-initiating pluripotency factor *PRDM14* (78) and multiple genes related to olfactory stimulus (*OR10H3*, *OR2T1*, *OR51V1*, and *UGT2A1*) (49). Taken together, these distal eQTL mappings suggest the ncRNAs have strong influences on gene expression of multiple pcGenes in both non-cancerous and tumor tissue.

#### Overlap of miRNA-pcGene pairs with target prediction databases

For in-silico validation, we queried TargetScan (79, 80), a database that curates computationally predicted RNA targets of miRNAs, for any miRNAs that our analysis detected to mediate distal-eQTLs of pcGenes. Out of 522 pairs of miRNAs and pcGenes across 72 unique miRNAs identified through our analysis, we found that 184 pairs were included in the TargetScan database (**Supplemental Table S3**). miRNA-pcGene pairs identified in our eQTL analysis are found in TargetScan at an enrichment ratio of 8.2 (95% CI: [6.68, 10.09]), compared to the universe of all miRNA-target pairs in TargetScan (approximately 3.56 million) and roughly 159,000 miRNA-target pairs in TargetScan for the 72 miRNA families identified. A majority of these miRNAs (82% of miRNAs detected in eQTLs) are conserved only across humans and mice but well-annotated. Though this intersection with TargetScan does not implicate the miRNA in distal regulation of the proposed pcGene, it provides some computational validation of this relationship using different methodology (sequence similarity vs. eQTL mapping).

### Distal-eGenes are enriched for tumorigenesis and cancer progression gene pathways

To assess enriched biological processes or pathways by sets of prioritized pcGenes (called distal-eGenes), we conducted gene ontology enrichments (81). Overall, compared to all expressed pcGenes in the transcriptome, distal-eGenes in non-cancerous tissue (combining breast and prostate) were significantly enriched (FDR-adjusted P < 0.05) for many relevant ontologies: immune processes, genes targeted by epigenetic regulation, microRNA targets in cancer, and oxidoreductase activity. In comparison, distal-eGenes detected in tumor tissue showed enrichments mainly for chemical and sensory receptors and intermediate filament cytoskeleton (Supplemental Figure S3-4). Comparing tissue-prioritized distal-eGenes (breast-specific or prostate-specific pcGenes to the protein-coding transcriptome), we found distal-eGenes enrichments detected in prostate tissue for immune pathway ontologies, including the multiple activations of immune cells, and known tumorigenic pathways, like the JAK-STAT cascade and PI3K-Akt signaling (82, 83). We did not detect any significant enrichments for the breast-specific distal-eGenes (Supplemental Figure S5).

We also conducted comparisons of distal-eGenes between non-cancerous and tumor state in both breast and prostate tissue (Figure 3). We find that, compared to distal-eGenes prioritized in breast tumors, non-cancerous breast distal-eGenes were enriched for cytokine and leukocyte production, response, and function, as well as membrane transport and binding. We observed similar enrichments when we compared non-cancerous prostate distal-eGenes to those from prostate tumors, with additional cell death and morphogenesis ontologies enrichments. In contrast, across both breast and prostate tissue, tumor-specific distal-eGenes, compared to non-cancerous distal-eGenes, mainly showed enrichments for olfactory and chemical stimulus response, intermediate filament cytoskeleton localization, and epidermis development. These ontologies are consistent with cancer progression, as olfactory receptors have been validated as prognostic biomarkers in prostate cancers and are overexpressed in more aggressive breast tumors (48,49,84,85), more aggressive breast cancers are enriched for genes that influence epidermal growth (86) and cytoskeletal dysregulation is key to cancer cell invasion, progression, and metastasis (87–89).

**Figure 3:**
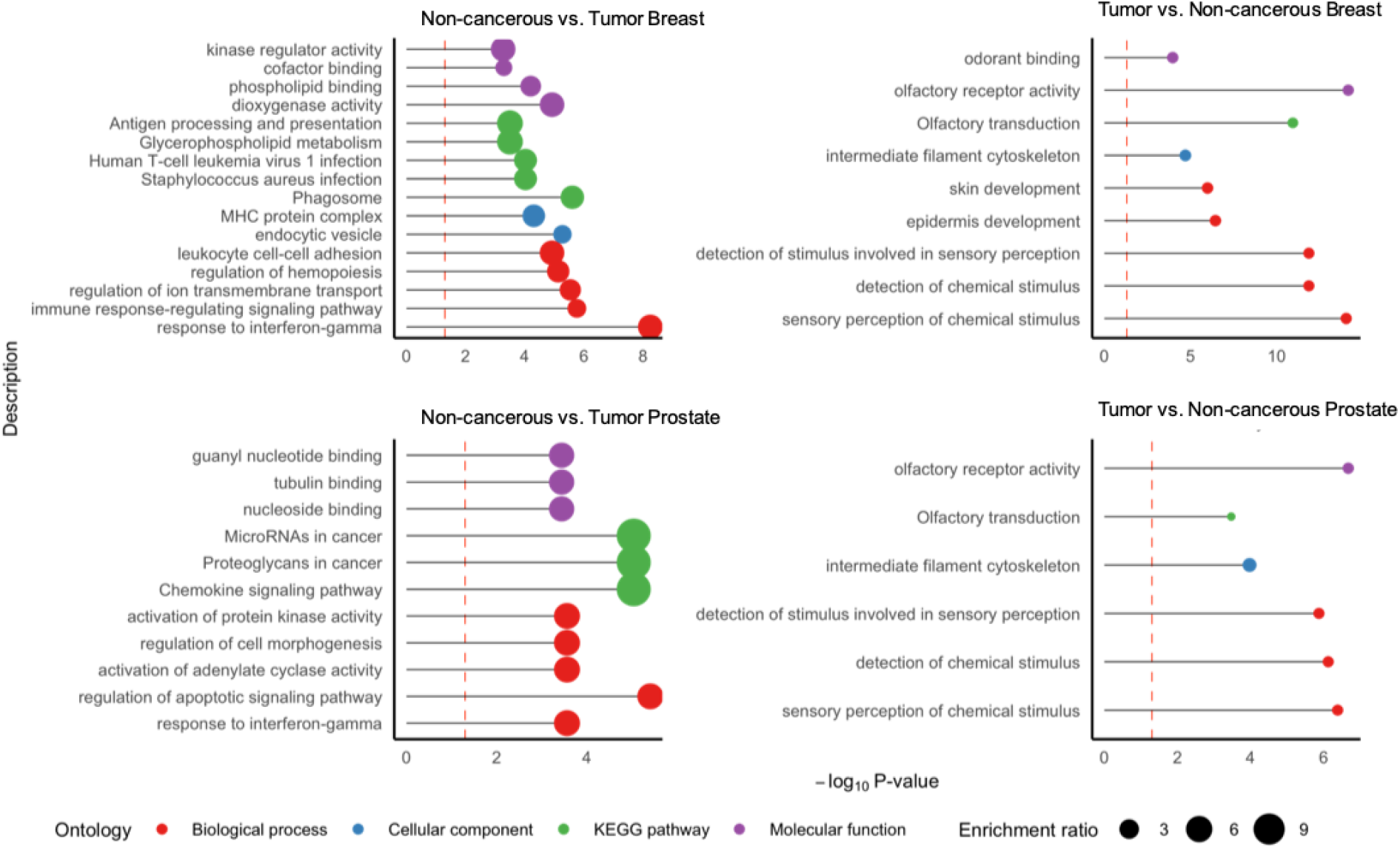
Over-represented ontologies for tissue-specific eGenes. -log_10_ P-value of enrichment (X-axis) of over-represented gene sets (Y-axis), with point sized by enrichment ratio and colored by ontology category. Here, for a tissue, we compare the set of pcGenes from healthy or tumor state to the universe of all pcGenes for tissue across both healthy and tumor states.

### Distal-eQTLs overlap with genetic signal for breast and prostate cancer risk

Lastly, we integrated these eQTL results with GWAS summary statistics for overall prostate and overall and molecular subtype-specific breast cancer risk (1, 2). A total of 84 detected ncRNAs are within 0.5 Mb of a GWAS SNP at P < 5 x 10^-8^, the majority for overall prostate and breast cancer risk, which have the largest GWAS sample sizes (Figure 4A**, Supplemental Table S1-2**). Among pairs of ncRNAs and pcGenes where the ncRNA is 0.5 Mb from a GWAS SNP for either breast or prostate cancer risk, we identified 30 pairs (15 for overall breast cancer and 10 for LumA breast cancer, five for prostate cancer) where at least one of the ncRNA local-eQTL or pcGene distal-eQTL colocalized with the GWAS signal at the locus with PP.H4 ≥ 0.75 (Supplemental Figure S6, **Supplemental Table S4**). In total, we detected 10 independent overall or LumA-specific breast cancer risk GWAS signals and one independent prostate cancer risk GWAS signal may be, in part, explained by distal genetics effects on pcGenes mediated by local-ncRNAs.

**Figure 4:**
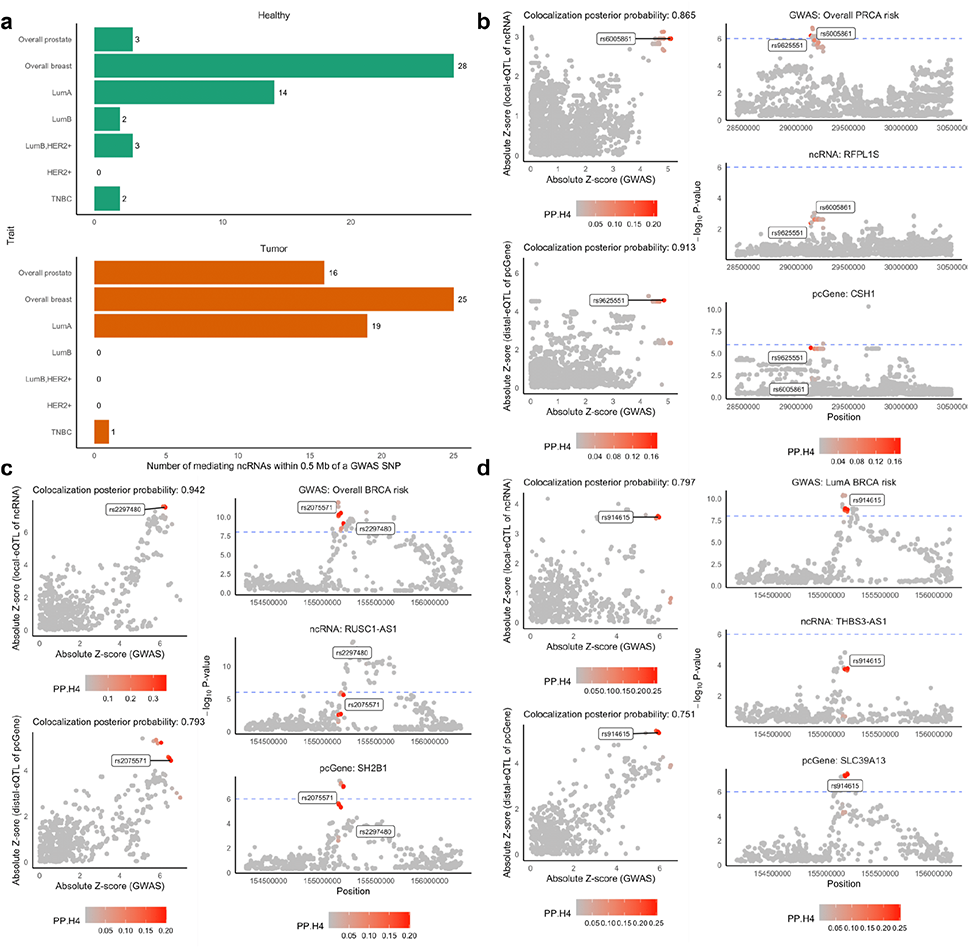
Colocalization of local-eQTLs of ncRNA and distal-eQTLs of pcGene with GWAS. **(a)** Barplot of numbers of mediating ncRNAs within 1 Megabase of a GWAS SNP (X-axis) from cancers (Y-axis). **(b-d)** Colocalization results for example ncRNAs and pcGenes, with phenotype in the GWAS, ncRNA, and pcGene provided. Left panel shows scatterplot of absolute Z-scores of GWAS (X-axis) and eQTL associations (Y-axis) with points colored by posterior probability of colocalization (PP.H4). Right panel shows a Manhattan plot of GWAS (top), ncRNA local-eQTL (middle), and pcGene distal-eQTL (bottom) signal, colored by PP.H4.

We found that distal-eQTLs of *CSH1* mediated by *RFPL1S* strongly colocalize with GWAS signal for prostate cancer risk. (PP.H4 = 0.913) (Figure 4B). *CSH1* codes for a somatotropin hormone with paracrine signaling functions promoting cell division and growth in glands (90). Using non-cancerous breast tissue eQTLs, we found strong colocalization with overall breast cancer risk with local-eQTLs of *RUSC1-AS1* and distal-eQTLs of *SH2B1* (Figure 4C). Two different SNPs carried the largest posterior probability of colocalization for the local-(rs2297480) and distal-eQTLs (rs2075571), which are in moderate, yet statistically significant, linkage disequilibrium (D’ = 0.51, R^2^ = 0.131, P < 0.001). *RUSC1-AS1* has shown evidenced silencing of genes through epigenetic signaling and is correlated with breast cancer progression (91, 92). Additionally, *SH2B1* is involved with cytokine signaling in cell proliferation and migration (93). We also found strong colocalization with LumA-specific breast cancer risk with local-eQTLs of *THBS3-AS1* and distal-eQTLs of *SLC39A13* in non-cancerous breast tissue. The same SNP showed the maximum posterior probability for colocalization with both the local- and distal-eQTL signal. Though the ncRNA has not been implicated in cancer risk or progression, *SLC39A13* facilitates metastasis in ovarian cancer by activating the Src/FAK signaling pathway (94).

For overall breast cancer risk, we also detect strong colocalization between eQTLs and GWAS signals at the 17q21.31 locus. This locus houses a large, common inversion polymorphism associated with breast and ovarian cancer prognosis (95–97), as well as widespread associations with multiple phenotypes (98–101). In particular, we found that the ncRNA *KANSL1-AS1* potentially mediates distal-eQTLs of multiple pcGenes. All but one of these pcGenes are on Chromosome 17, near the end of the 17q21.31 region; the last pcGene we detected is *TXNRD3* at 3q21.3. We were interested in disentangling effects of the H2 inversion on gene expression and breast cancer risk, with the proposed causal diagram presented in Figure 5A. First, we estimated haplotypes of the H2 inversion in GTEx (102). We found that *KANSL1-AS1* and the associated pcGenes near 17q21.31 all have significant associations with the H2 inversion (Figure 5B). Next, we reran mediation analysis for SNPs local to *KANSL-AS1* and these detected pcGenes. After accounting for H2, the distal mediation signal of *KANSL-AS1* is attenuated for all pcGenes except *TXNRD3*, suggesting that the inversion may be driving a significant portion of this signal. These analyses support our proposed causal model, that the genetics of the H2 inversion affect expression of the local ncRNA KANSL-AS1 and the distal pcGenes at the end of the inversion, which induces the eQTL associations (Figure 5A).

**Figure 5:**
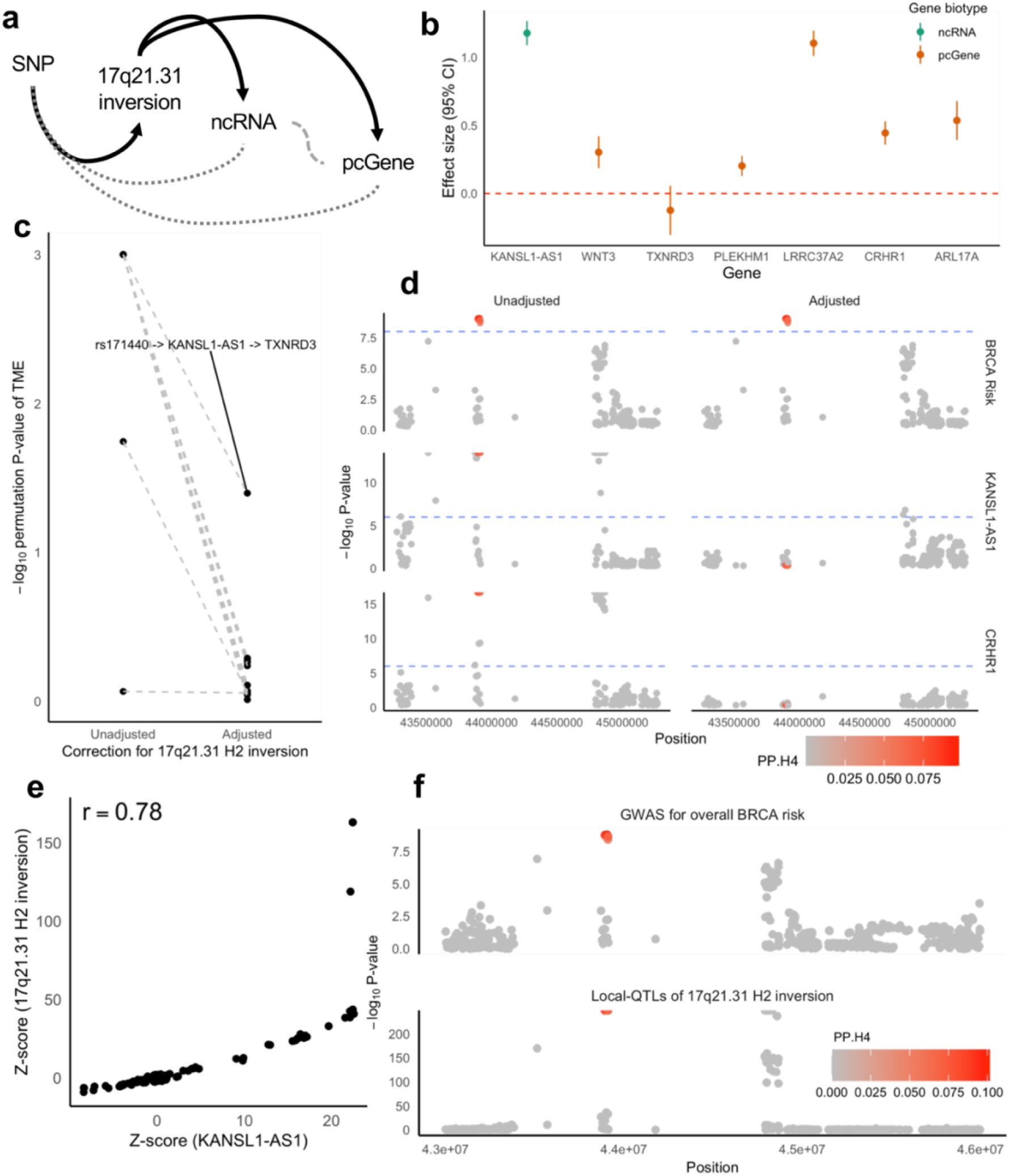
Impact of H2 inversion on eQTLs in the 17q21.31 locus. **(a)** Causal diagram of genetic effects in 17q21.31, where strong effects of genetically-determined H2 inversion on ncRNA and pcGene (in black) induces the observed SNP-ncRNA and -pcGene associations (in grey). **(b)** Forest plot of effect sizes and 95% confidence interval (Y-axis) on H2 inversion on pcGenes or ncRNAs (X-axis). **(c)** Difference in −log10 permutation P-value (Y-axis) of total mediation effect of ncRNA on pcGene, with or without adjustment for H2 inversion (X-axis). **(d)** Manhattan plots of GWAS, local-eQTLs of KANSL-AS1, and distal-eQTLs of CRHR1, unadjusted (left) and adjusted (right) for H2 inversion, colored by per-SNP PP.H4. **(e)** Scatterplot of Z-score of local-eQTLs on KANSL1-AS1 (X-axis) against Z-score of local-QTLs of H2 inversion. **(f)** Manhattan plots of GWAS and local-QTLs of H2 inversion, colored by per-SNP PP.H4.

H2 inversion-adjusted colocalization analyses also support this model. A salient example is shown in Figure 5D, with Manhattan plots from colocalization analysis of overall breast cancer risk, local-eQTLs of *KANSL-AS1*, and distal-eQTLs of *CRHR1*. Colocalization analysis unadjusted for H2, showed nearly-perfect colocalization for both eQTL signals with GWAS (PP.H4 > 0.98 for both eQTLs), but after adjustments, the eQTL signals are completely removed. In fact, the standardized effect sizes for local-eQTLs of *KANSL1-AS1* showed strong correlation with the standardized effect sizes for local-QTLs of the H2 inversion (Figure 5E), and the local-QTL signal for the H2 inversion strongly colocalized (PP.H4 = 0.99) with the GWAS signal in the locus (Figure 5F). These results illustrate that the structural inversion in the locus likely accounts for both *cis*-genetic control of *KANSL1-AS1* and the associated pcGenes further downstream on Chromosome 17. Furthermore, we emphasize further examination of the H2 inversion and other structural variants in the locus for its impact on local and distal gene expression, as well as cancer susceptibility, especially to elucidate if SNPs with widespread local or distal associations with gene expression are affected by confounding due to structural variants or other aberrations.

We also studied the cancer risk associations of ncRNAs with predicted large transcriptomic effects using a genetically-regulated expression (GReX) approach (**Methods**). At P < 2.5 x 10^-6^ and permutation P < 0.05, we identify 33 ncRNA-level associations (Figure 6, **Supplemental Table S5**), predominantly for overall and subtype-specific breast cancer risk. Only one ncRNA, *SDHAP2*, showed an association with overall prostate cancer risk (Figure 6); as we restrict to ncRNAs mediating distal-eQTLs and not necessarily in known prostate cancer GWAS loci, we do not recover any ncRNAs detected by Guo et al’s analysis of long-ncRNAs in prostate cancer (103). A couple ncRNAs prioritized in associations of breast cancer risk using ncRNA GReX in non-cancerous breast tissues have been previously noted. Common non-synonymous SNPs in *HCG9* have previously been implicated in breast cancer GWAS (104). In addition, *CEROX1*, a cataloged post-transcriptional regulator of mitochondrial catalytic activity, has been implicated in distal alterations of metabolic pathways in breast cancers (14, 105). ncRNAs prioritized in breast tumor GReX-associations with breast cancer risk mainly included micro- and snoRNAs. Of these, *miR-519d* has been shown to suppress breast cancer cell growth by targeting distal molecular features(106, 107). GReX associations prioritize these ncRNAs for further investigation of their functional effects in breast and prostate tissue.

**Figure 6:**
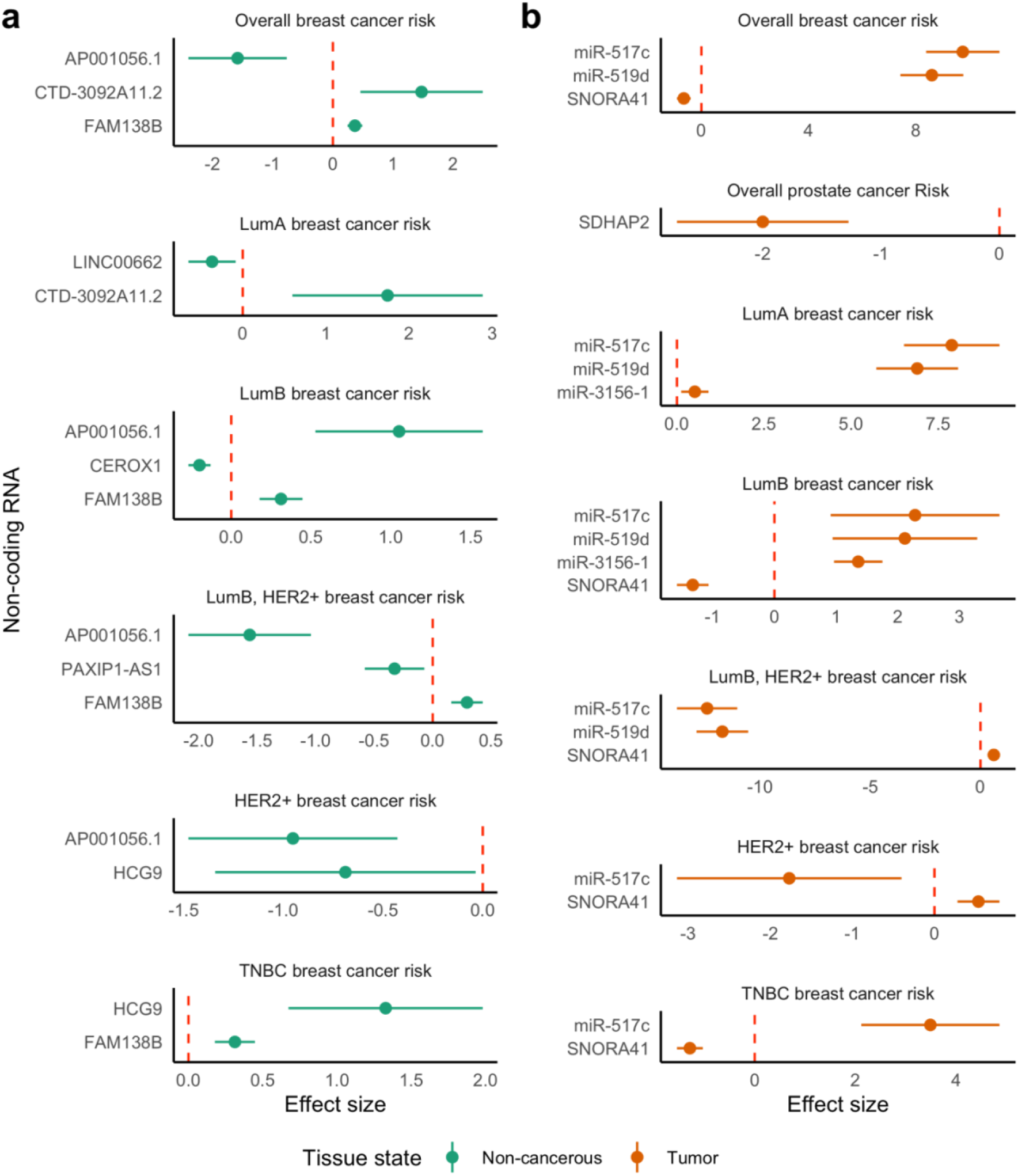
GReX associations with cancer risk for ncRNAs mediating multiple distal-eQTLs of pcGenes. Forest plot of effect size and confidence intervals at significance level of P = 2.5 x 10^-6^ (Y-axis) of GReX-associations with overall and subtype-specific cancer risk across ncRNAs that showed significant mediation of multiple distal-eQTLs of distinct pcGenes (X-axis) in non-cancerous **(a)** and tumor **(b)** tissue states.

## DISCUSSION

In this work, we systematically identify distal-eQTLs of pcGenes that are mediated by ncRNAs in non-cancerous and tumor breast and prostate tissue. We then show that many of these ncRNA-mediated distal-eQTLs of pcGenes overlap with GWAS signals for breast and prostate cancer, both with known GWAS loci and novel genetic loci undetected by GWAS. Taken together, our results suggest that distal genetic effects on pcGenes mediated by ncRNAs may be a common mechanism underlying genetic signals and ncRNAs have a widespread role as distal transcriptional regulators in prostate and breast tissue. We observed more distal ncRNA-pcGene directional associations in tumor than non-cancerous tissue, suggesting that tumor tissues have multiple activated gene regulatory networks with potential effects on disease pathogenesis or progression. Our results implicating distal interactions between ncRNAs and pcGenes are attractive for further in silico and experimental study.

We find many colocalized eQTL signals for ncRNAs and pcGenes in the 17q21.31 region, many of which have been prioritized by previous genetic association studies. For example, a transcriptome-wide association study (TWAS) of estrogen receptor subtype-specific breast cancer, identified ncRNA *KANSL-AS1* (108), which has also been associated with ovarian cancer (109). In our analysis, *KANSL-AS1* mediated multiple breast cancer-associated pcGenes further downstream on Chromosome 17 and colocalized almost perfectly with the GWAS association, suggesting widespread distal effects of *KANSL-AS1* (108,110–114). However, after accounting for the large and common H2 inversion in the 17q21.31 locus, the distal effects of *KANSL1-AS1* were largely attenuated, pointing to correlated effects of large structural aberrations on gene expression and disease etiology in this region (115–118). These results also suggest that structural aberrations, like the 17q21.31 H2 inversion, have large effects on gene expression, not only locally, but also distally. Our results also serve as a cautionary tale: future gene expression analyses must delineate the eQTL signal in a locus from chromosomal aberrations, especially when integrating with GWAS signals. Comprehensive analyses of the transcriptomic effects of genomic aberrations in tumor tissue are needed.

We conclude with limitations of our study. First, we rely on Ensembl annotations of gene biotypes to define our sets of ncRNAs and pcGenes. These annotations may be incomplete, and accordingly, we may have ignored multiple non-coding transcripts (119). Next, as our local- and distal-eQTL signals are the same cohor, we were unable to use multi-trait colocalization methods, like *moloc* or *Primo* (120, 121). A more flexible framework that allows for shared molecular QTL signal could be developed to fully interrogate the mediated-distal QTL signal, in the context of complex trait etiology. Next, we do not account for copy number variation or structural variation to disentangle these potentially disparate signals. Future studies should consider corrections for inversions, translocations, or genomic imbalances. Lastly, due to limited sample sizes in TCGA, we could not assess the effects of molecular subtype heterogeneity on eQTL mapping in tumor tissue. Previously, subtype-specific genetic architecture of gene expression regulation has been suggested by previous studies (5,122,123), with some distal genetic associations detected for genes that are highly predictive of molecular subtypes (124). Robust subtype-specific analyses in breast cancer can be informative for both subtype-specific risk and outcomes.

Our study provides evidence supporting *trans*-acting regulation by ncRNAs as a potential biological mechanism relevant to breast and prostate cancer etiology. Particularly, our results emphasize that larger samples of tissue- and tumor-specific transcriptomics datasets need to be collected to study often ignored transcripts and explore more complex regulatory hypotheses to interpret GWAS risk loci for cancer.

## Data Availability

GTEx v8 genotype, expression, and covariate data were obtained through dbGAP Study Accession phs000424.v8.p2. TCGA genotype were obtained through dbGAP Study Accession phs000178.v11.p8 and expression and covariate data was obtained from the Broad GDAC Firehose repository (https://gdac.broadinstitute.org). Prostate cancer risk summary statistics were obtained from the Prostate Cancer Association Group to Investigate Cancer Associated Alterations in the Genome (PRACTICAL) Consortium: http://practical.icr.ac.uk/blog/wp-content/uploads/uploadedfiles/oncoarray/MetaSummaryData/meta_v3_onco_euro_overall_ChrAll_1_release.zip. Breast cancer risk summary statistics were obtained from the Breast Cancer Association Consortium (BCAC): https://bcac.ccge.medschl.cam.ac.uk/bcacdata/oncoarray/oncoarray-and-combined-summary-result/gwas-summary-associations-breast-cancer-risk-2020/. Sample code for this analysis are available at https://github.com/ColetheStatistician/ncRNAInBreastCancer/.

http://practical.icr.ac.uk/blog/wp-content/uploads/uploadedfiles/oncoarray/MetaSummaryData/meta_v3_onco_euro_overall_ChrAll_1_release.zip

https://bcac.ccge.medschl.cam.ac.uk/bcacdata/oncoarray/oncoarray-and-combined-summary-result/gwas-summary-associations-breast-cancer-risk-2020/

https://github.com/ColetheStatistician/ncRNAInBreastCancer/

https://www.ncbi.nlm.nih.gov/projects/gap/cgi-bin/study.cgi?study_id=phs000424.v8.p2

https://www.ncbi.nlm.nih.gov/projects/gap/cgi-bin/study.cgi?study_id=phs000178.v11.p8

## ACKNOWLEDGEMENTS

We thank Nicholas Mancuso, Harold Pimentel, Mike Love, Achal Patel, and Kangcheng Hou for engaging conversation during the research process. We also thank the UCLA Bruins-in-Genomics Summer Program for the research opportunities.

The Prostate cancer genome-wide association analyses are supported by the Canadian Institutes of Health Research, European Commission’s Seventh Framework Programme grant agreement n° 223175 (HEALTH-F2-2009-223175), Cancer Research UK Grants C5047/A7357, C1287/A10118, C1287/A16563, C5047/A3354, C5047/A10692, C16913/A6135, and The National Institute of Health (NIH) Cancer Post-Cancer GWAS initiative grant: No. 1 U19 CA 148537-01 (the GAME-ON initiative).

Genotyping of the OncoArray was funded by the US National Institutes of Health (NIH) [U19 CA 148537 for ELucidating Loci Involved in Prostate cancer SuscEptibility (ELLIPSE) project and X01HG007492 to the Center for Inherited Disease Research (CIDR) under contract number HHSN268201200008I] and by Cancer Research UK grant A8197/A16565. Additional analytic support was provided by NIH NCI U01 CA188392 (PI: Schumacher).

We would also like to thank the following for funding support: The Institute of Cancer Research and The Everyman Campaign, The Prostate Cancer Research Foundation, Prostate Research Campaign UK (now PCUK), The Orchid Cancer Appeal, Rosetrees Trust, The National Cancer Research Network UK, The National Cancer Research Institute (NCRI) UK. We are grateful for support of NIHR funding to the NIHR Biomedical Research Centre at The Institute of Cancer Research and The Royal Marsden NHS Foundation Trust.

The breast cancer genome-wide association analyses for BCAC and CIMBA were supported by Cancer Research UK (C1287/A10118, C1287/A16563, C1287/A10710, C12292/A20861, C12292/A11174, C1281/A12014, C5047/A8384, C5047/A15007, C5047/A10692, C8197/A16565), The National Institutes of Health (CA128978, X01HG007492-the DRIVE consortium), the PERSPECTIVE project supported by the Government of Canada through Genome Canada and the Canadian Institutes of Health Research (grant GPH-129344) and the Ministère de l’Économie, Science et Innovation du Québec through Genome Québec and the PSRSIIRI-701 grant, the Quebec Breast Cancer Foundation, the European Community’s Seventh Framework Programme under grant agreement n° 223175 (HEALTH-F2-2009-223175) (COGS), the European Union’s Horizon 2020 Research and Innovation Programme (634935 and 633784), the Post-Cancer GWAS initiative (U19 CA148537, CA148065 and CA148112 - the GAME-ON initiative), the Department of Defence (W81XWH-10-1-0341), the Canadian Institutes of Health Research (CIHR) for the CIHR Team in Familial Risks of Breast Cancer (CRN-87521), the Komen Foundation for the Cure, the Breast Cancer Research Foundation and the Ovarian Cancer Research Fund. All studies and funders are listed in Zhang H et al (Nat Genet, 2020).

## FUNDING

TS was supported by the National Institute of Neurological Disorders and Stroke of the National Institutes of Health under Award Number T32NS048004. This research was supported by the National Institute of Mental Health of the National Institutes of Health under Award number 5R01MH115676-04. AG was partially supported by R01 CA227237. SL was partially supported by NIH award R01 CA194393. BP were partially supported by NIH awards R01 HG009120, R01 MH115676, R01 CA251555, R01 AI153827, R01 HG006399, R01 CA244670, U01 HG011715. The content is solely the responsibility of the authors and does not necessarily represent the official views of the National Institutes of Health.

## AUTHOR INFORMATION

### Contributions

AB conceived the study. AB, TS, and PL developed the statistical approaches, performed the analysis, and drafted the paper. AB, TS, and BP provided insight in methodological approaches and analysis. BP provided data resources. AB and BP supervised the study. All authors approved and edited the final manuscript.

### Corresponding author

Correspondence to Arjun Bhattacharya

## ETHICS DECLARATION

### Ethics approval and consent to participate

This study was approved by the Office of Human Research Ethics at the University of California, Los Angeles, and written informed consent was obtained from each participant. All experimental methods abided by the Helsinki Declaration.

### Consent for publication

Not applicable.

### Competing interests

The authors declare that they have no competing interests.

## SUPPLEMENTAL FIGURES

**Supplemental Figure S1:**
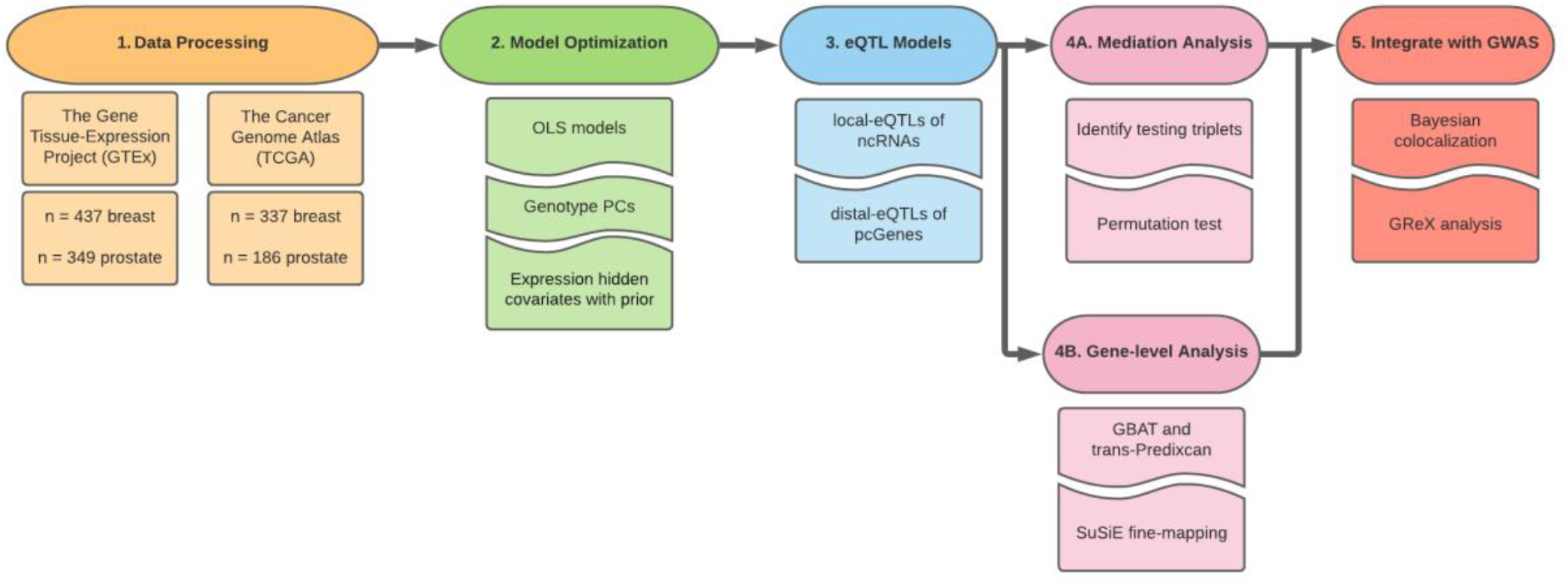
Analysis scheme. We analyze genetic and transcriptomic data from non-cancerous breast and prostate tissue from GTEx and breast and prostate tumors from TCGA. We optimize eQTL mapping using ordinary least squares regression for numbers of genotype principal components and expression hidden covariates with prior to optimize eQTL discovery. We then conduct a genome-wide local and distal eQTL analysis using the optimized set of covariates. Next, we conduct mediation analysis or gene-based association testing to identify distal-eQTLs of pcGenes that are mediated by local-eQTLs of ncRNAs. Lastly, we integrate eQTLs results with GWAS using colocalization and analysis of genetically-regulated expression.

**Supplemental Figure S2:**
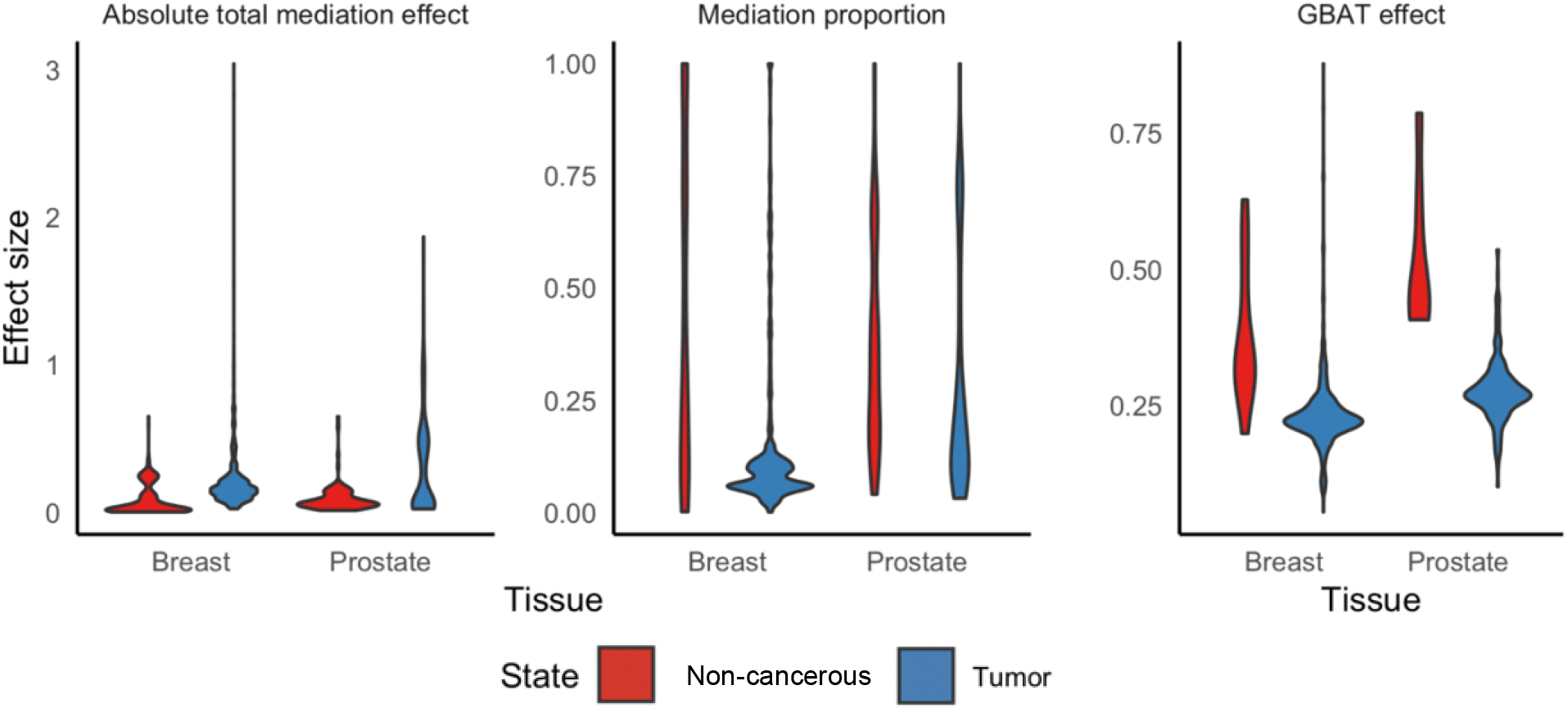
Distribution of total mediation effects, mediation proportions, and gene-level distal effect sizes in healthy and tumor breast and prostate tissue.

**Supplemental Figure S3:**
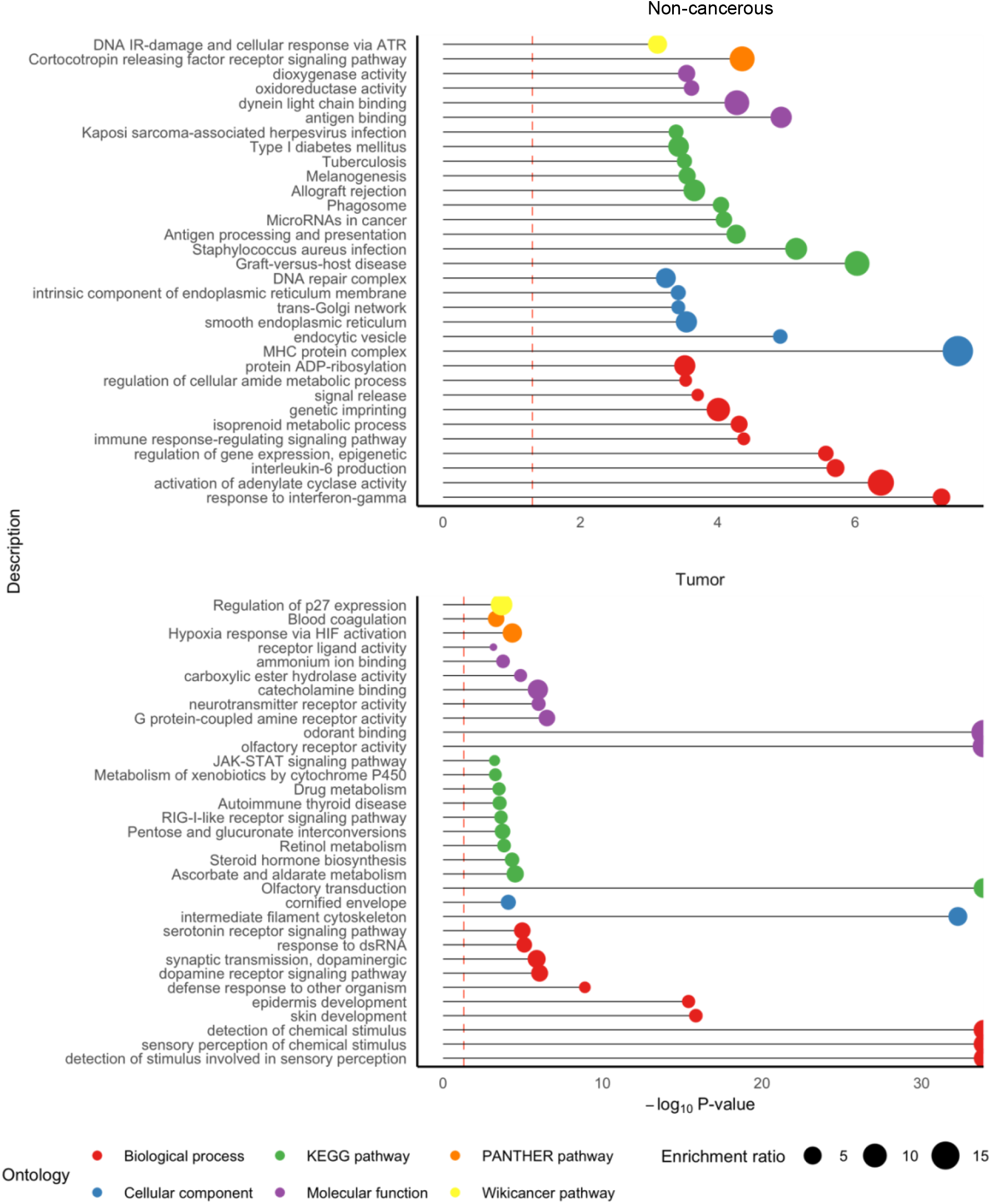
Over-represented ontologies for healthy- or tumor-specific eGenes, compared to all protein-coding genes in the transcriptome. -log_10_ P-value of enrichment (X-axis) of over-represented gene sets (Y-axis), with point sized by enrichment ratio and colored by ontology category. Here, combining pcGenes with a distal genetic association across breast and prostate tissue, we compare the set of pcGenes from healthy or tumor state to the universe of all pcGenes in the transcriptome.

**Supplemental Figure S4:**
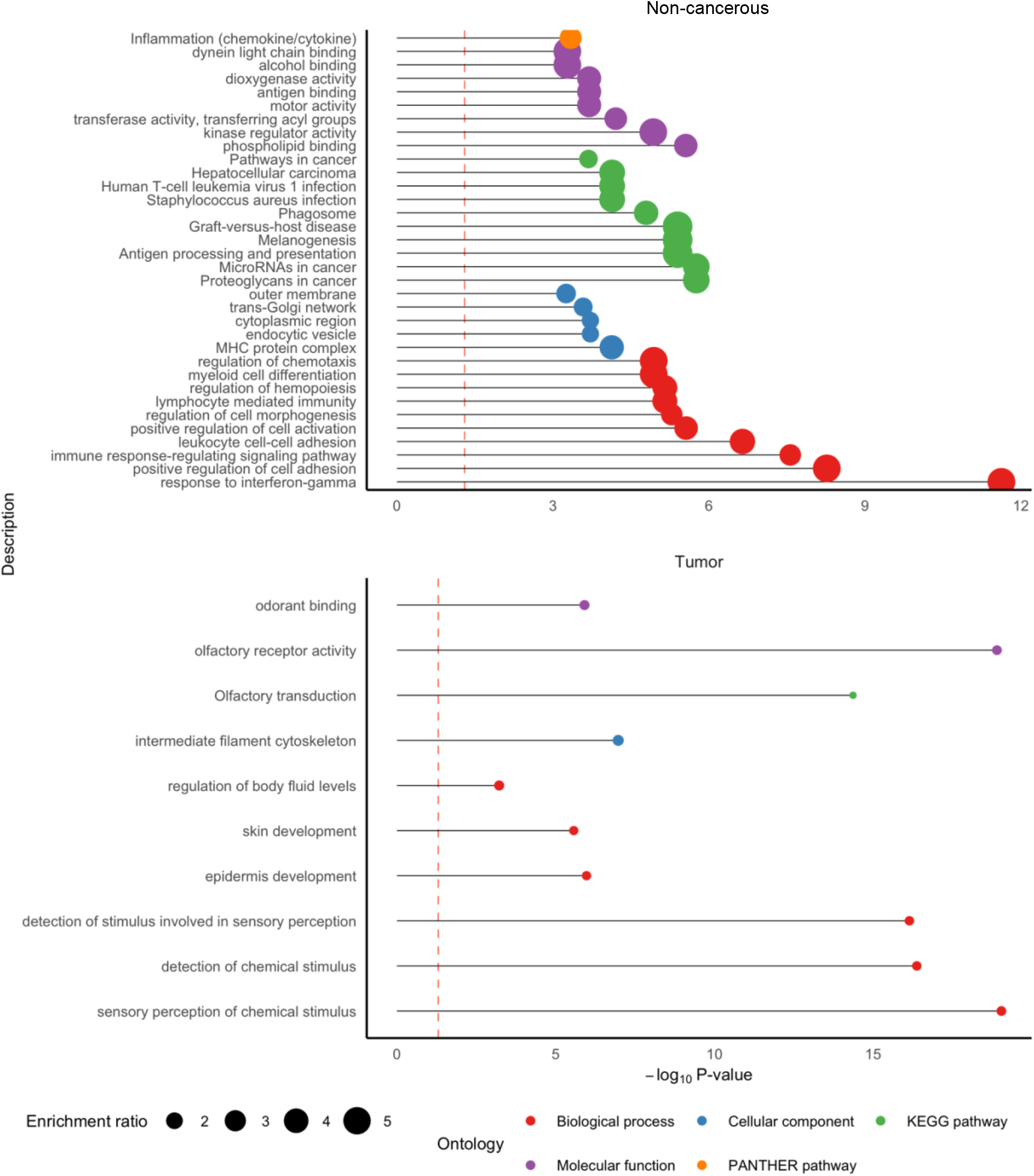
Over-represented ontologies for healthy or tumor state-specific eGenes, compared to all protein-coding genes detected in distal-QTL mapping. -log_10_ P-value of enrichment (X-axis) of over-represented gene sets (Y-axis), with point sized by enrichment ratio and colored by ontology category. Here, combining pcGenes with a distal genetic association across breast and prostate tissue, we compare the set of pcGenes detected for breast or prostate and compare to the universe of all pcGenes detected across breast and prostate. No enrichments at P < 0.05 were detected for breast-specific pcGenes.

**Supplemental Figure S5:**
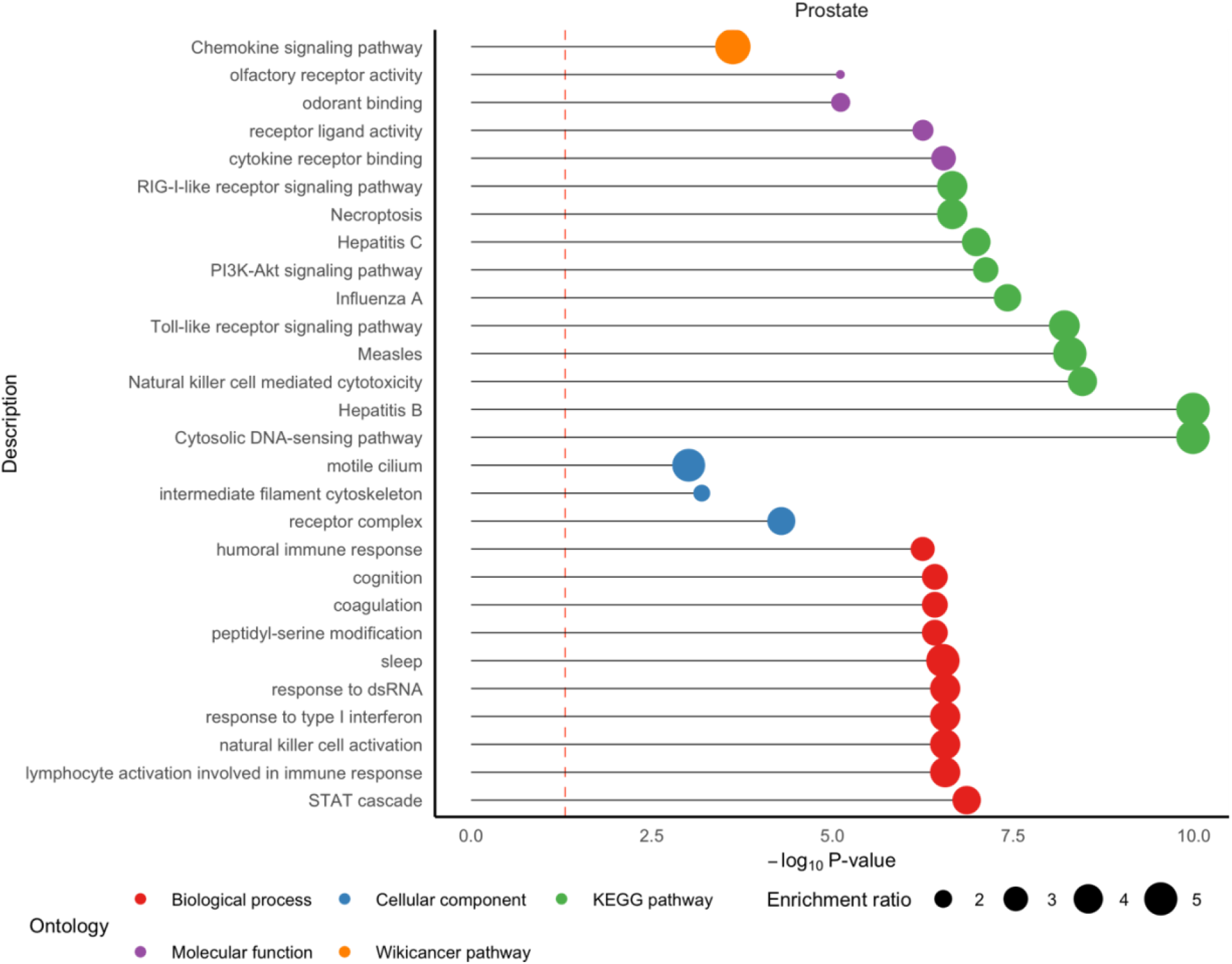
Over-represented ontologies for breast or prostate-specific eGenes, compared to all protein-coding genes detected in distal-QTL mapping. -log_10_ P-value of enrichment (X-axis) of over-represented gene sets (Y-axis), with point sized by enrichment ratio and colored by ontology category. Here, combining pcGenes with a distal genetic association across healthy and tumor state, we compare the set of pcGenes detected for breast or prostate and compare to the universe of all pcGenes detected across breast and prostate. No enrichments at P < 0.05 were detected for breast-specific pcGenes.

**Supplemental Figure S6:**
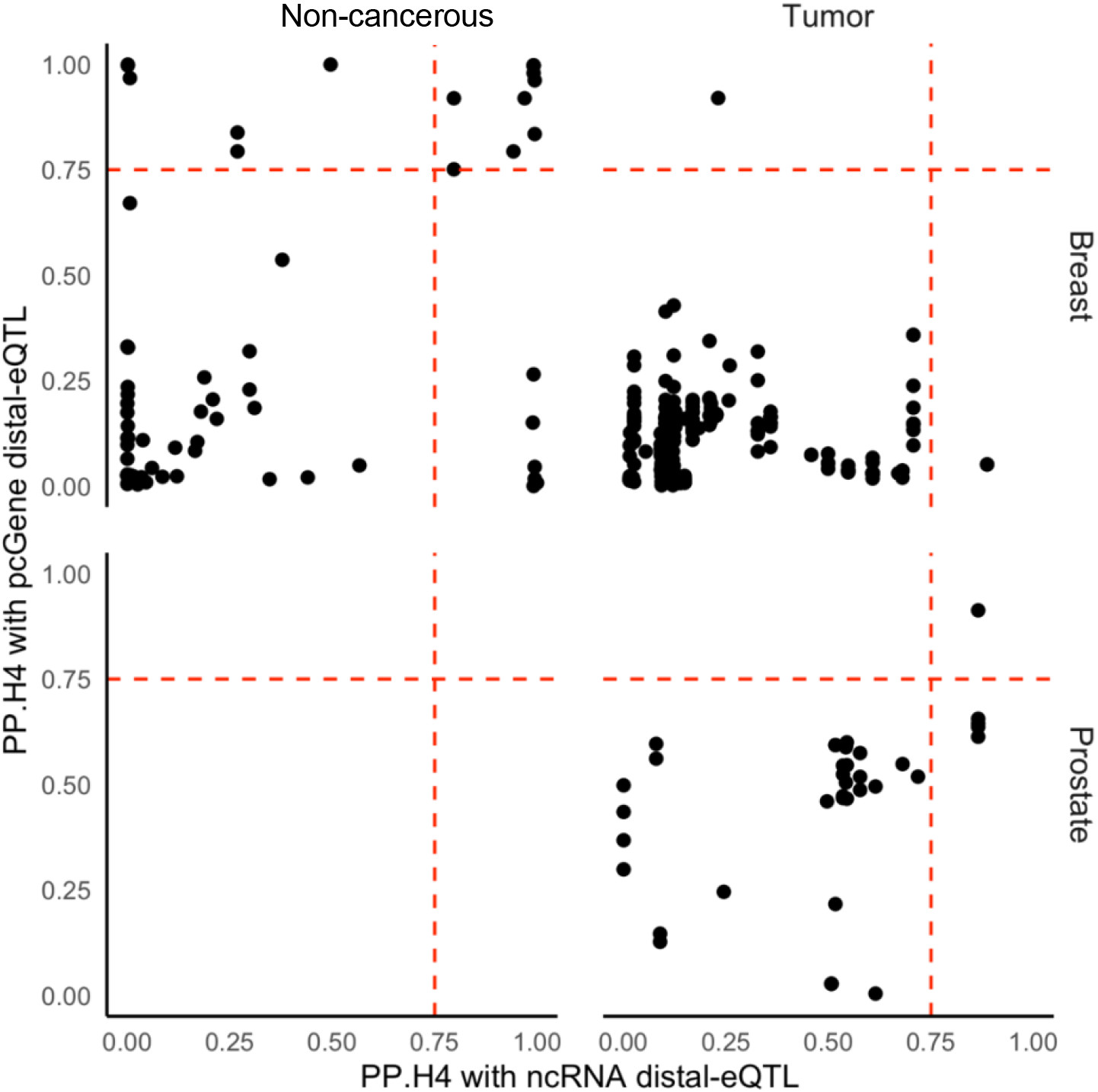
Scatterplot of posterior probability of colocalization for local- and distal-eQTLs with cancer risk GWAS. Red lines show PP.H4 = .75.

### SUPPLEMENTAL TABLE LEGENDS

**Table S1:** P*r*ioritized *ncRNA-mediated distal-eQTLs of pcGenes in healthy and tumor prostate tissue.* We provide the tissue state (healthy or tumor), SNP, ncRNA, pcGene, effect size, P-value, method of detection (mediation analysis or GBAT), and closest GWAS risk SNP and P-value.

**Table S2:** P*r*ioritized *ncRNA-mediated distal-eQTLs of pcGenes in healthy and tumor breast tissue.* We provide the tissue state (healthy or tumor), SNP, ncRNA, pcGene, effect size, P-value, method of detection (mediation analysis or GBAT), and closest GWAS risk SNP and P-value.

**Table S3:** *In-silico validation of miRNA-pcGene pairs using TargetScan.* For miRNAs detected to mediated distal-eQTLs of pcGenes, we list miRNAs shown to target the pcGene using TargetScan.

**Table S4:** C*o*localization *results for ncRNA-mediated distal-eQTLs with GWAS signal of cancer risk.* We provide the trait, tissue (breast or prostate), tissue state (healthy or tumor), ncRNA and its location, pcGene and its location, posterior probabilities of colocalization between the ncRNA and pcGene with the GWAS signal, and the colocalized SNP for each signal.

**Table S5:** *GReX analysis results for ncRNAs and cancer risk.* We provide the trait, tissue (breast or prostate), tissue state (healthy or tumor), ncRNA and its location, effect size, standard error, and Z-score of association.

## Notes

### Competing Interest Statement

The authors have declared no competing interest.

### Author Declarations

This study was approved by the Office of Human Research Ethics at the University of California, Los Angeles, and written informed consent was obtained from each participant.

### Summary of Updates

Tighten language

